# Gene expression signatures identify biologically and clinically distinct tuberculosis endotypes

**DOI:** 10.1101/2020.05.13.20100776

**Authors:** Andrew R. DiNardo, Tanmay Gandhi, Jan Heyckendorf, Sandra L. Grimm, Kimal Rajapakshe, Tomoki Nishiguchi, Maja Reimann, Jaqueline Kahari, Qiniso Dlamini, Christoph Lange, Torsten Goldmann, Sebastian Marwitz, DZIF-TB cohort study group, Abhimanyu, Jeffrey D. Cirillo, Stefan HE Kaufmann, Mihai G. Netea, Reinout van Crevel, Anna M. Mandalakas, Cristian Coarfa, Members of the DZIF-TB cohort study group

## Abstract

**Background:** *In vitro*, animal model, and clinical evidence suggests that tuberculosis is not a monomorphic disease, and that host response to tuberculosis is protean with multiple distinct molecular pathways and pathologies (endotypes). We applied unbiased clustering to identify separate tuberculosis endotypes with classifiable gene expression patterns and clinical outcomes.

**Methods:** A cohort comprised of microarray gene expression data from microbiologically confirmed tuberculosis patients were used to identify putative endotypes. One microarray cohort with longitudinal clinical outcomes was reserved for validation, as was one RNA-seq cohorts. Finally, a separate cohort of tuberculosis patients with functional immune results was evaluated to clarify stimulated from unstimulated immune responses.

**Results:** A discovery cohort, including 435 tuberculosis patients and 533 asymptomatic controls, identified two tuberculosis endotypes. Tuberculosis patient endotype A is characterized by increased expression of genes related to inflammation and immunity and decreased metabolism and proliferation; in contrast, endotype B increased activity of metabolism and proliferation pathways. An independent RNA-seq validation cohort, including 118 tuberculosis patients and 179 controls, validated the discovery results. Gene expression signatures for treatment failure were elevated in endotype A in the discovery cohort, and a separate validation cohort confirmed that endotype A patients had slower time to culture conversion, and a reduced incidence of cure. These observations suggest that endotypes reflect functional immunity, supported by the observation that tuberculosis patients with a hyperinflammatory endotype have less responsive cytokine production upon stimulation.

**Conclusion:** These findings provide evidence that metabolic and immune profiling could inform optimization of endotype-specific host-directed therapies for tuberculosis.

## INTRODUCTION

Host-directed therapies (HDT) could improve the efficacy, shorten the duration of treatment regimens, or ameliorate tuberculosis (TB)-induced lung pathology. The fields of asthma, COPD, and most cancers have identified biological endotypes, distinct cellular pathology. Consequently, treatment for these diseases depends on the specific pathways that are disturbed, with endotype-specific therapies improving clinical outcomes[1]. For example, asthma endotypes can generally be divided into neutrophilic versus eosinophilic disease, with the former being more responsive to macrolide therapy and the latter being corticosteroid responsive [1]. Leprosy is also treated based on immune endotypes, with the cell-mediated and paucibacillary form requiring less antibiotics for a shorter duration, while the anergic and multibacillary endotype requires more antibiotics for a longer duration. No similar categorization system is available to guide TB HDT.

Studies have identified incongruous immune responses that can lead to TB[2-7]. To further test the premise that there is not a single stereotypical immune response to TB, we sought to provide evidence for the diversity of host responses during TB. Mycobacterial immunity requires a balanced, well-regulated response from multiple cell types. For example, immune control of *Mycobacterium tuberculosis* (*Mtb)* requires tightly regulated IFN-γ and TNF□ with decreased IFN-γ or TNF□ resulting in decreased *Mtb* intracellular killing capacity[4], but with exuberant IFN-γ or TNF□ inducing macrophage and tissue necrosis with extracellular *Mtb* survival[2, 3]. To better characterize TB endotypes, we implemented an unbiased clustering of publicly available gene expression data, then validated the results using two external cohorts, eventually identifying a hyperinflammatory TB endotype associated with worse clinical outcomes.

## METHODS

### Study inclusion

A systematic review and meta-analysis were implemented according to PRISMA guidelines (described in detail in supplemental methods). Publicly available data was identified using PubMed and the NCBI Gene Expression Omnibus (GEO) repository. Studies without microbiologic confirmation, without description of the methods of microbiologic confirmation, or without evaluation of whole blood were excluded. Twenty-two gene expression studies from whole blood were identified that included participants with microbiologically confirmed pulmonary TB. Studies or datasets that included only cases without controls or evaluated fewer than 10,000 genes were excluded. Studies using microarray were used for a discovery cohort and RNA-seq datasets for a validation cohort (Supplemental Table 1). One microarray cohort (GSE147689, GSE147691) was reserved for validation because it contained longitudinal clinical outcomes (Table 1; Supplemental Table 1).Normalization, processing, clustering, and development of a gene classifier is described in full in the supplemental methods. Clinical outcomes for this cohort[8] were defined using the TBNET criteria: cure is defined as culture-negative at 6 months, with no positive cultures thereafter and no disease relapse within a year after treatment completion. Treatment failure was based on a positive culture 6 months after treatment initiation or disease relapse within one year after treatment completion.

**Table 1:** Epidemiologic characteristics of the German and Romanian validation cohort. (MDR = multi-drug resistance; TTP = time to positivity; BMI = body mass index; TCC = time to culture conversion). (Not all data was available for all participants.) Mann-Whitney test performed for continuous variables and one-sided chi-squared evaluated differences in populations. Not all data was available for all participants.

|  | All | Endotype A | Endotype B | p-value* |
| --- | --- | --- | --- | --- |
| Median age | 37.95 | 39.78 | 37.95 | 0.6948 |
| % Male | 62.2% (56/90) | 62.5% (30/48) | 61.9% (26/42) | 0.4768 |
| Median BMI | 20.75 | 19.49 | 21.28 | 0.0009 |
| Current or previous smoker | 52.0% (51/98) | 46.2% (24/52) | 58.7% (27/46) | 0.1074 |
| Cavitary Disease | 71.7% (66/92) | 76.5% (39/51) | 65.9% (27/41) | 0.1305 |
| MDR TB | 62.0% (75/121) | 54.7% (35/64) | 70.2% (40/57) | 0.0399 |
| Baseline TTP (days) | 17 | 14 | 21 | 0.0169 |
| Median TCC (days) | 54.0 | 64.5 | 33.5 | 0.0005 |
| Cure | 81.0% (51/63) | 74.4% (29/39) | 91.7% (22/24) | 0.0447 |
| Mortality | 6.3% (4/63) | 10.3% (4/39) | 0% (0/24) | 0.0525 |

MDR, multi-drug resistance; TTP, time to positivity; BMI, body mass index; TCC, time to culture conversion. (Not all data was available for all participants)
\*Mann-Whitney U test or one-sided Chi-squared (A worse than B) were used to determine significance, as appropriate.

### Immunology Validation Cohort

Multiplex ELISA (LegendPlex) was implemented with and without overnight mitogen stimulation in a cohort of pulmonary TB patients (n = 40) or their asymptomatic household contacts (n = 39) from Eswatini. The study protocol was reviewed by institutional review boards and all participation was voluntary and in concordance with the Declaration of Helsinki. TB patients were defined by both the presence of symptoms and microbiologic confirmation by culture or Gene Xpert. Twenty-two (55%) of the TB cases and nine (23%) of the household contacts were HIV co-infected, respectively.

### Statistics

Chi-squared test with a one-sided tail assessed incidence of clinical variables between endotypes. Rank-sum of the cytokines/chemokines from the ELISA validation was used to stratify TB patients. Differences between sub-groups were analyzed using Mann-Whitney rank sum test.

## RESULTS

### Systematic selection of TB patient cohorts with whole blood transcriptomic profiles

Seven studies profiled whole blood and included both cases and controls by microarray gene expression analysis, profiling 435 individuals with microbiologically confirmed TB and 533 healthy controls[9-15]. These 7 studies, comprising 12 datasets in NCBI GEO, were used for unbiased clustering to identify TB endotypes, based on 12,468 commonly evaluated genes (Figure 1; Supplemental Table 1). Two additional studies with both cases and controls used RNA-seq transcriptome profiling and included 118 TB patients and 179 controls[16, 17]. These studies were reserved for creation of an RNA-seq validation cohort (Supplemental Table 1). A cohort from Germany and Romania including 121 TB patients and 14 healthy controls using microarrays was reserved as an additional external validation dataset because it included clinical outcome data (Table 1)[18]. A fourth cohort of TB patients and healthy controls with multiplex ELISA at baseline and phytohemagglutinin stimulated whole blood (Figure 1).

**Figure 1:**
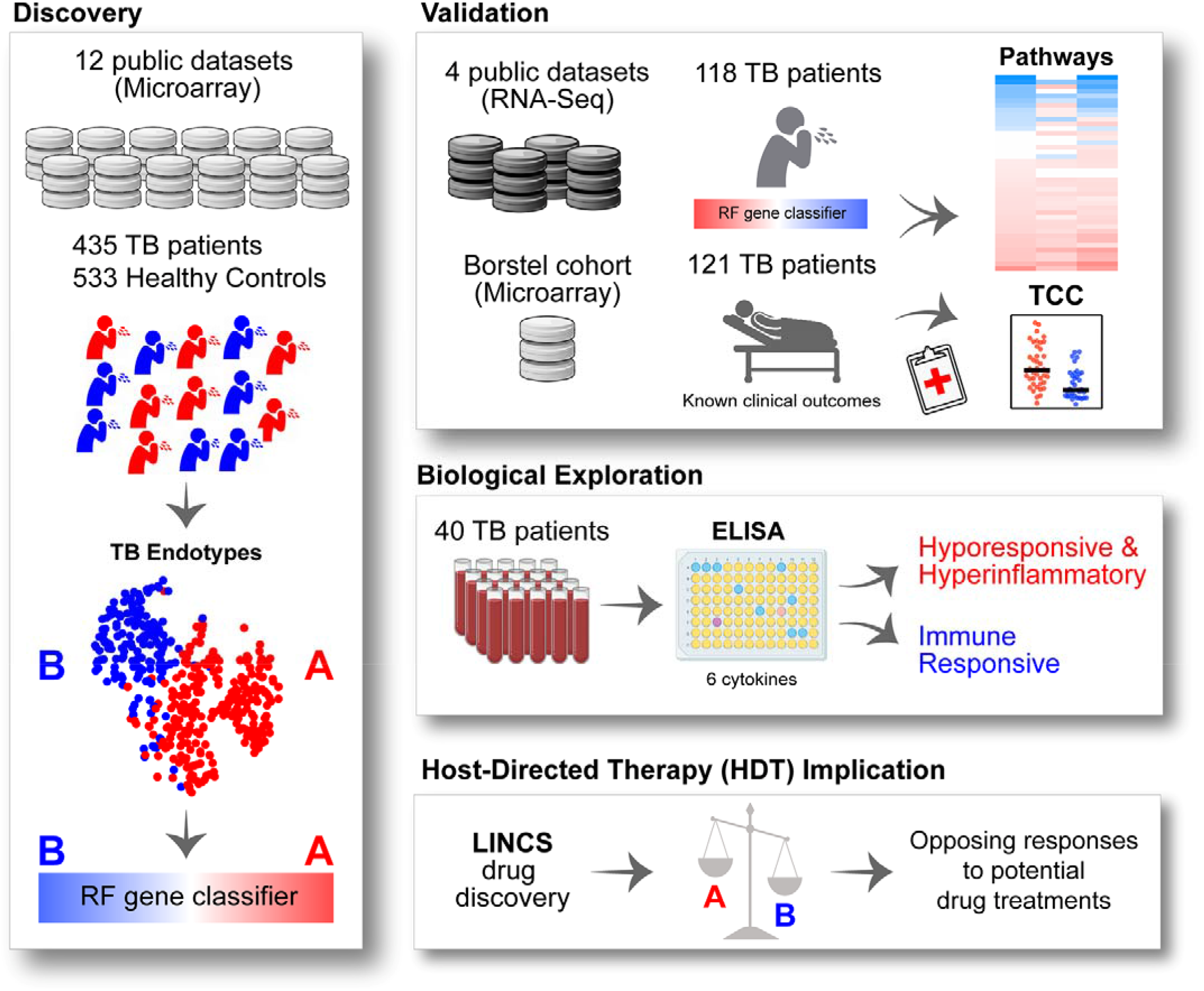
Overview of study. Using unbiased clustering of TB patients from 12 publicly available microarray datasets a Random Forest gene classifier was derived to predict TB Endotype A versus B. This was validated on an RNA-seq cohort compiled over 4 publicly available datasets and on a microarray patient cohort that included longitudinal clinical outcomes data. Immunological validation was evaluated by multiplex ELISA using a separate cohort from Eswatini. Finally, similarity to endotype gene signatures was used to assess and rank previously evaluated host directed therapy candidates.

### Identification and clinical data characterization of TB endotypes

To identify potential TB endotypes, unbiased clustering of the microarray transcriptome of 435 TB patients was performed (Figure 2A-C). Clustering was evaluated at various resolutions (Supplemental Figure 1) with final analysis being performed at resolution 0.4 due to limited discriminatory capacity at higher resolutions. At resolution 0.4, two distinct endotypes were identified. Endotype A consisted of 269 TB patients (54.8%), and endotype B included 166 TB patients (45.1%). Patients from each country and each study were well distributed in both endotype clusters (Figure 2C; Supplemental Table 1). In the discovery cohort, only two datasets included individuals co-infected with HIV (GSE37250, and GSE39939; Supplemental Table 1). The 108 TB patients living with HIV clustered into endotype A and endotype B, with 57 and 51 in each respectively. Only one study included children under 15 years of age (GSE39939), with 23 children clustering into endotype A and 12 clustering into endotype B (Supplemental Table 1).

**Figure 2:**
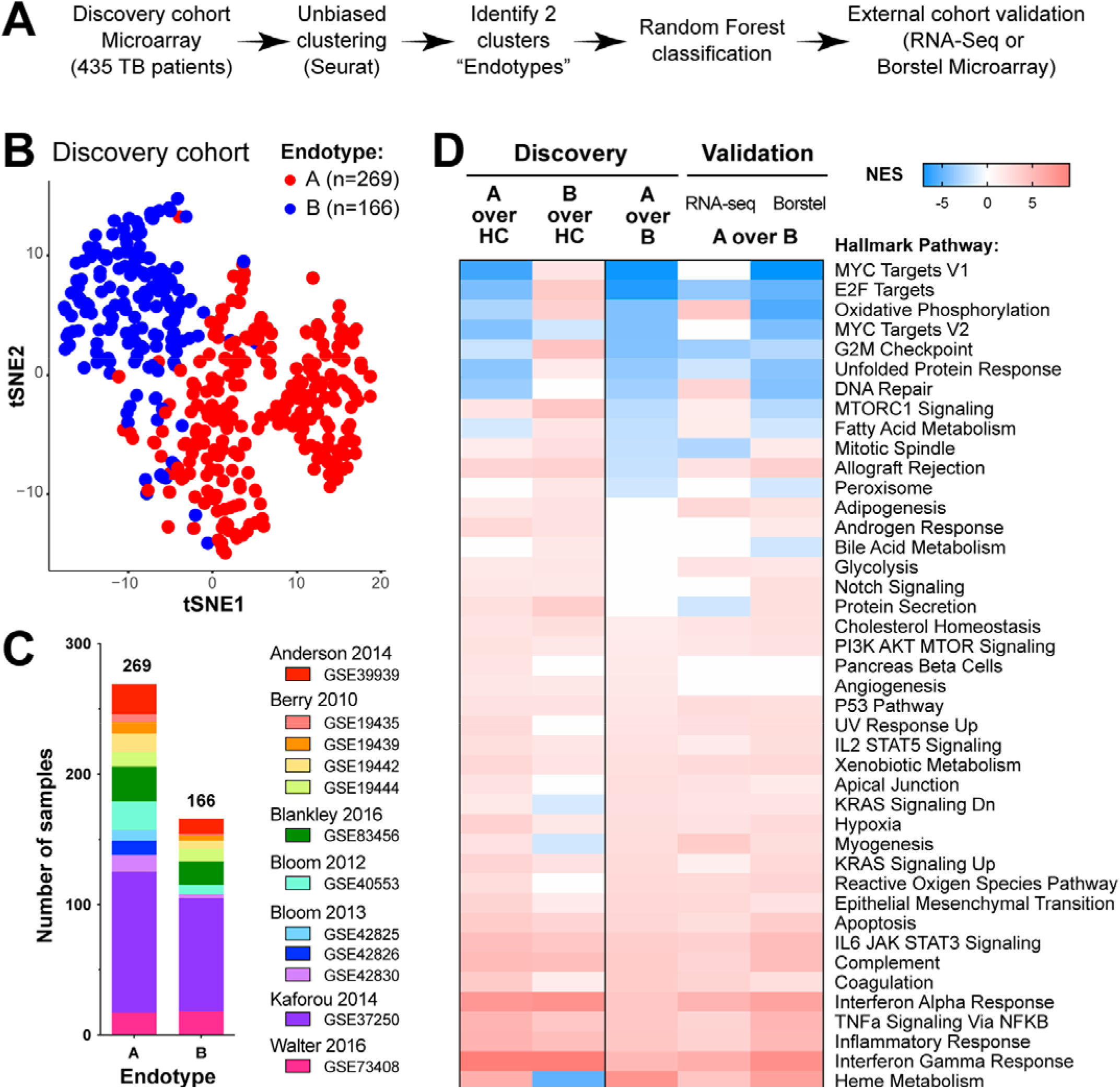
Unbiased clustering identifies unique TB endotypes. (A) Unbiased clustering was implemented on discovery cohort of 7 studies, identifying two major endotypes; next, a random forest gene classifier was developed and applied to two external validation datasets. (B) Network-based unbiased clustering using the Louvain method identifies 2 major endotypes of TB. (C) Distribution of individual studies into Endotype A or B. (D) TB endotypes were compared to healthy controls and against each other, then pathway enrichment via Gene Set Enrichment Analysis (GSEA) was carried out against the Hallmark pathway compendium.

Using the random forest machine learning algorithm, a classifier was derived to categorize endotype A vs endotype B based on the discovery cohort. Genes were ranked by their individual classification score, then accuracy of a random forest gene classifier was evaluated across a spectrum of genes using the measured classification error (Supplementary methods; Supplemental Figure 2A). A 40-gene classifier had both a low misclassification rate and was comprised of a low gene count. We applied the endotype classifier to a validation cohort comprised of 118 TB patients and 179 controls from two previously published RNA-seq studies (Supplemental Table 1). In the RNA-seq validation cohort, 65 TB samples classified as endotype A (55%) and 53 as endotype B (45%; Supplemental Table 1). We computed pathway enrichment using GSEA between each of the predicted endotypes and the control samples, and for the comparison between the endotypes. Random forest classification captured the significant divergent gene expression enrichments for immune related pathways (Figure 2D, Supplemental Figure 2B). For one study, microarray profiles (GSE19442 and GSE19444) were used in the discovery dataset whereas RNA-seq profiles (GSE107991) for the same samples were also used in the RNA-Seq validation cohort. Our classifier achieved good concordance between the microarray discovery cohort endotypes and the predicted RNA-seq endotypes (Supplemental Figure 3). Similar to the discovery cohort, endotype A enriched for inflammation, IFN-γ signaling, TNF□, and heme metabolism, while endotype B enriched for pathways related to cellular proliferation including E2F, G2M and mitosis (Figure 2D). Compared to healthy controls, gene targets of the transcription factors E2F, ELK1, and NRF1 were increased in endotype B, but were decreased in endotype A (Supplemental Table 3; Supplemental Figure 4). The endotypes were evaluated against six previously published scores that identified risk of treatment failure, with two of the scores also identifying risk for TB disease severity (Supplemental methods). Endotype A exhibited higher scores for risk of treatment failure and more severe disease compared to healthy controls (Figure 3).

**Figure 3:**
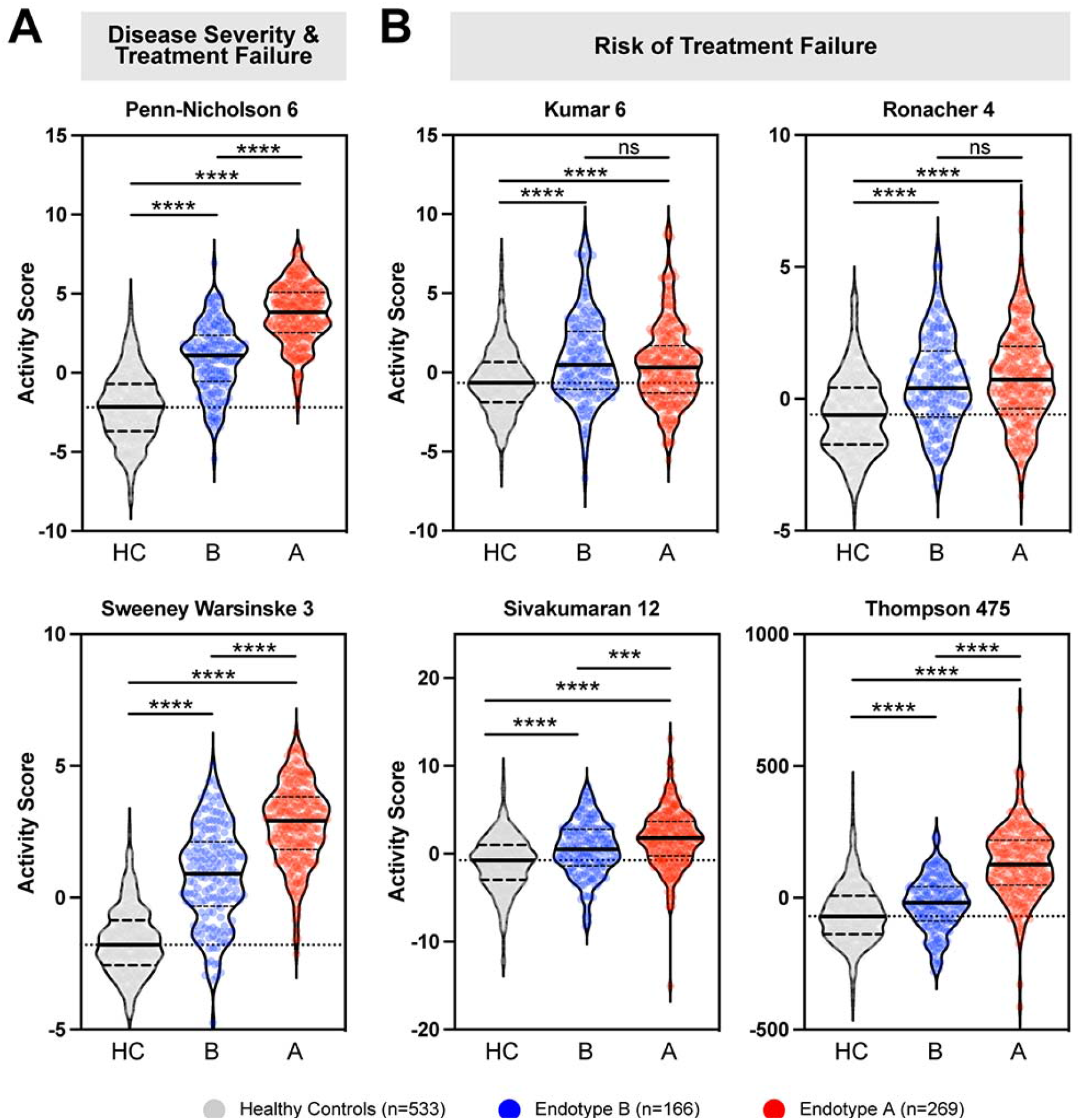
Evaluation of TB risk signatures over the TB endotypes. Previous studies identified gene signatures associated with A) disease severity and treatment failure or with B) risk of treatment failure. Activity scores were computed for each risk signature across healthy controls (HC, grey) and Endotype A (A, red) and Endotype B (B, blue). Mann-Whitney scores with **** p < 0.0001, *** p < 0.0002, ns = non-significant. Solid line at median, dashed lines at interquartile range, dotted line across at median of HC.

### Differential clinical outcome between TB endotypes

The classifier was applied to a cohort of TB patients assayed using microarray from a prospective, multicenter trial in Germany and Romania[18]. This cohort contained information on baseline bacillary burden, time to culture conversion, and clinical outcomes defined by the TBNET criteria (Table 1). Of 121 TB patients, 64 classified as endotype A (53%), while 57 classified as endotype B (47%). Similar to the RNA-seq validation cohort, endotype A and B demonstrated distinct enriched pathway profiles (Figure 2D, Supplemental Figure 2B). Based on the increased predicted treatment failure and disease severity signatures (Figure 3), we hypothesized that endotype A patients will display worse clinical outcome compared to the endotype B patients. Whereas endotype A had a lower rate of multi-drug resistance (54.7% vs 70.2%), Endotype A had slightly increased rates of cavitary disease (endotype A 76.5% vs. endotype B 65.9%, p= 0.1305), higher initial bacterial load (14 versus 21 days; p= 0.0169), slower times to culture clearance (64.6 days versus 33.5 days, p = 0.0005; Figure 4A; Table 1) and decreased rates of cure outcomes (74.4% versus 91.7%, p = 0.0447; Figure 4B; Table 1). All deaths occurred in endotype A (11.7% versus 0%, p = 0.0525).

**Figure 4:**
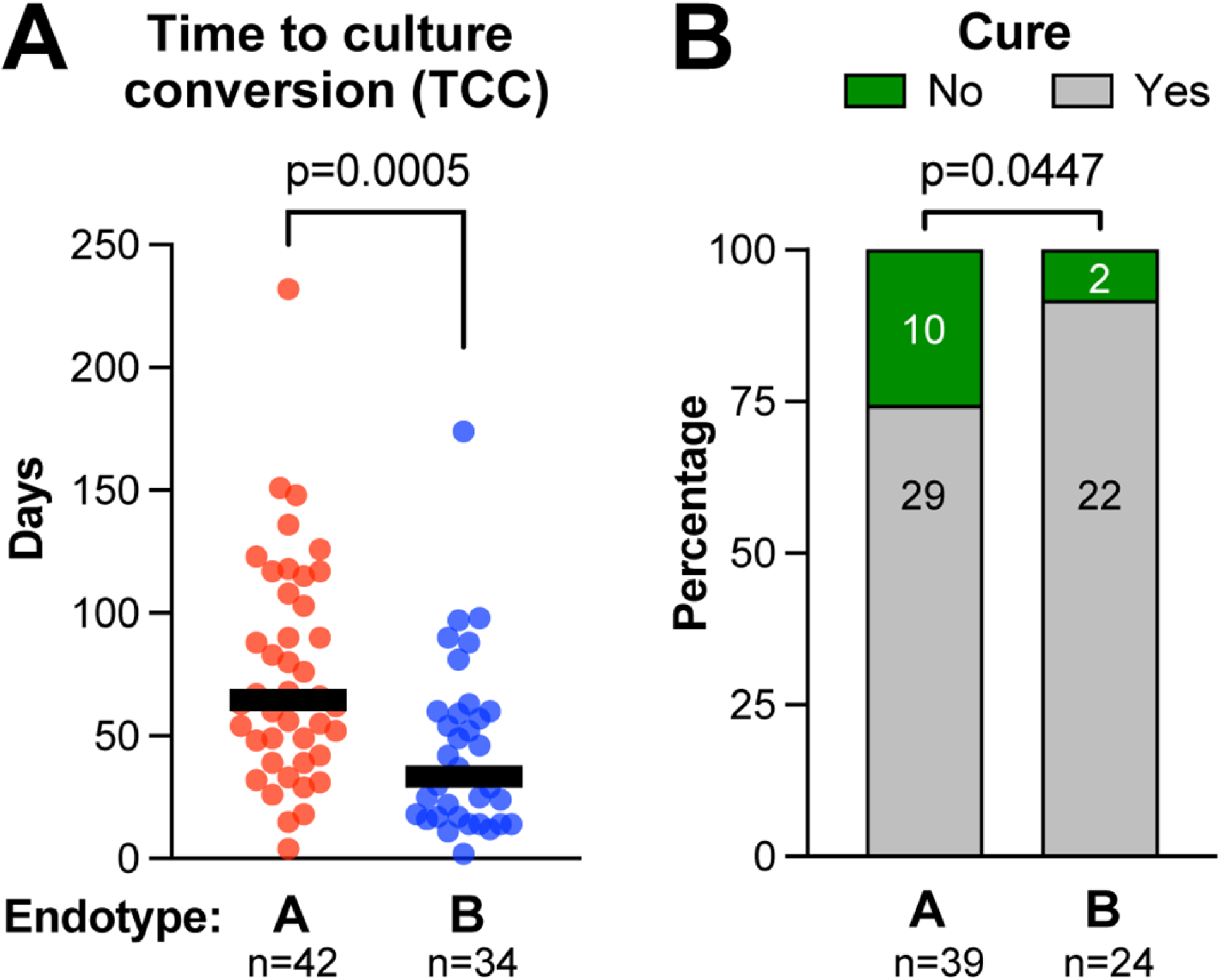
Endotype evaluation of TB clinical outcomes. Using the clinical annotations of the Borstel TB cohort, outcome differences between endotypes and association of pathway scores with outcomes were evaluated. (A) Time to culture conversion (TCC) in TB patients identified as endotype A or B (p = 0.0005 by Mann Whitney). (B) Rates of diseases cure in TB patients identified as endotype A or B (p = 0.0447 by one-sided Chi-squared test).

### Characterizing transcriptome trajectories across endotypes

To understand the relationships between controls and endotype A and B, we employed Cell Trajectory (also termed pseudotime trajectory) based on transcriptomic profiles. The trajectory score increased from healthy controls to endotype B to endotype A (Figure 5A); this result led to a search for specific molecular properties that follow the predicted disease trajectory. We computed normalized pathway activity scores for each patient in the discovery cohort. The results demonstrate that, in general, pathways related to inflammation and immunity increase in a monotonic manner from healthy controls to endotype B to endotype A (Figure 5B). Upon acute infection, increased glycolysis, the tricarboxylic acid cycle (TCA) and one-carbon metabolism provide metabolites requisite to fuel cellular proliferation; however, if infection is chronic, cells become metabolically exhausted and proliferation decreases[19-22]. Compared to healthy controls, endotype B has increased expression of pathways related to metabolism and proliferation (oxidative phosphorylation, electron transport chain (ETC), and G2M; Figure 5C-D; Supplemental Table 3). In contrast, endotype A patients exhibit decreased activity scores for pathways related to cellular proliferation (G2M, MYC and E2F; p < 0.0001) and metabolism (oxidative phosphorylation, the TCA cycle and the ETC; p < 0.001; Figure 5C-D).

**Figure 5:**
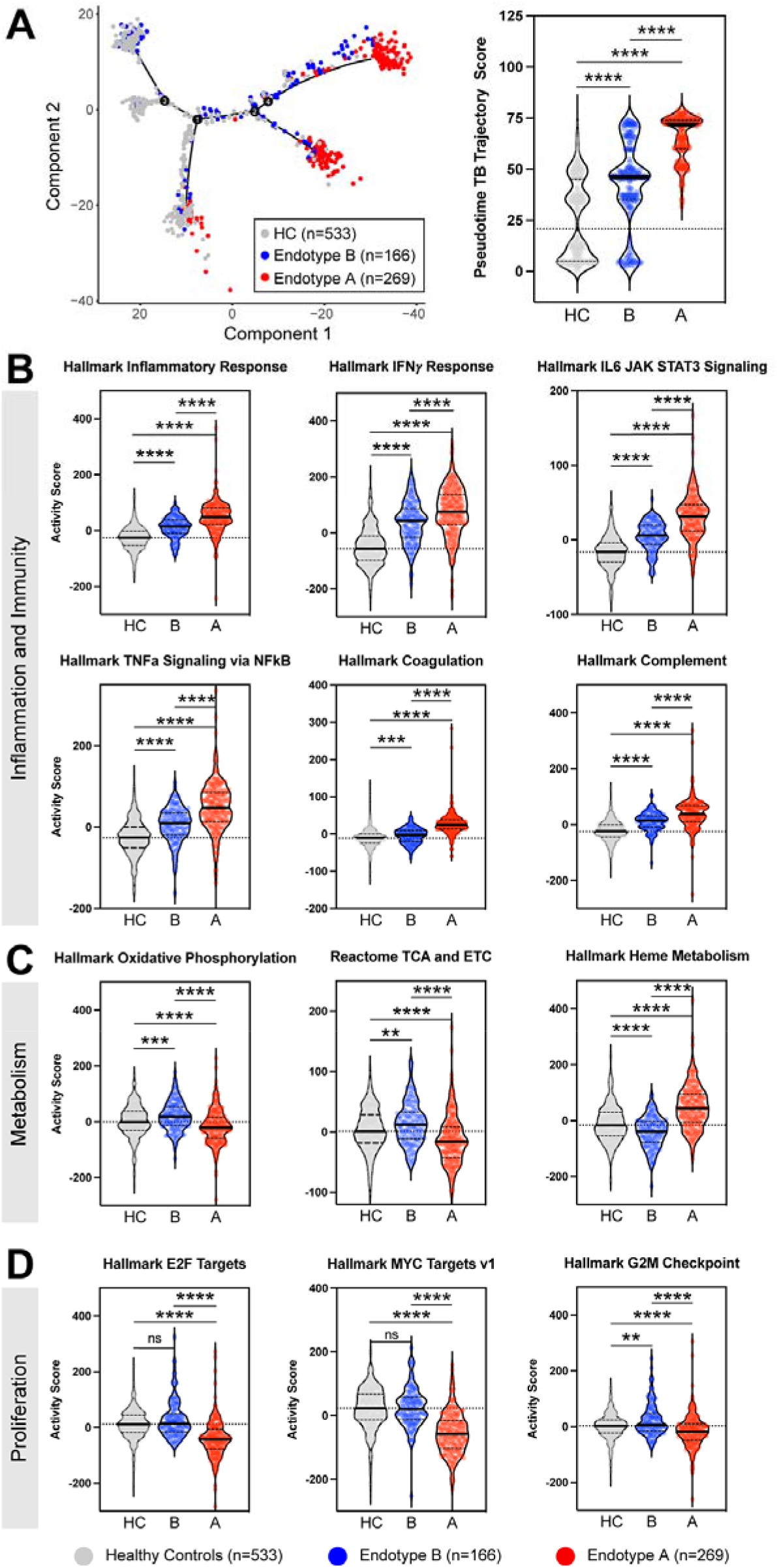
TB endotypes display distinct immune and metabolic gene expression activity scores. (A) Pseudotime TB trajectory score. Pathway activity scores were evaluated between healthy controls (HC, grey), endotype B (B, blue) and endotype A (A, red). Inflammation and immunity pathways (B), Metabolic pathways (C), and Proliferation pathways (D). Mann-Whitney test was used; **** p < 0.0001, *** p < 0.0002, ** p < 0.0021, ns = non-significant. Solid line at median, dashed lines indicate interquartile range, dotted line across HC median. Specific gene changes are available in Supplemental Table 2.

### Hyperinflammatory, hyporesponsive TB endotype

At the gene transcription level, TB endotype A is characterized by increased inflammation and increased interferon, TNF□, and IL-6 signaling in non-stimulated blood (Figure 2D). To evaluate the functional response upon stimulation, an independent cohort from Eswatini with mitogen-stimulated whole blood samples was analyzed by ELISA. Rank-sum analysis was implemented to stratify the patients into immune-responsive versus less-responsive groups based on their response to stimulation with mitogen (phytohemagglutinin) (Figure 6A-B). The two TB patient sub-groups were compared to healthy controls. The hypo-responsive group demonstrated a baseline hyperinflammatory condition, similar to endotype A, but decreased capacity to up-regulate IFN-γ, TNF□, IL-1β, IL-6, CXCL9, and CXCL10 upon stimulation with mitogen (Figure 6C-6D, p < 0.007). The immune-responsive group was similar to the healthy controls with regard to the capacity to respond to stimulation. Among the 40 TB patients, there were 4 deaths, 3 in the hyperinflammatory, hypo-responsive group and 1 in the immune responsive group (χ^2^ p = 0.27).

**Figure 6:**
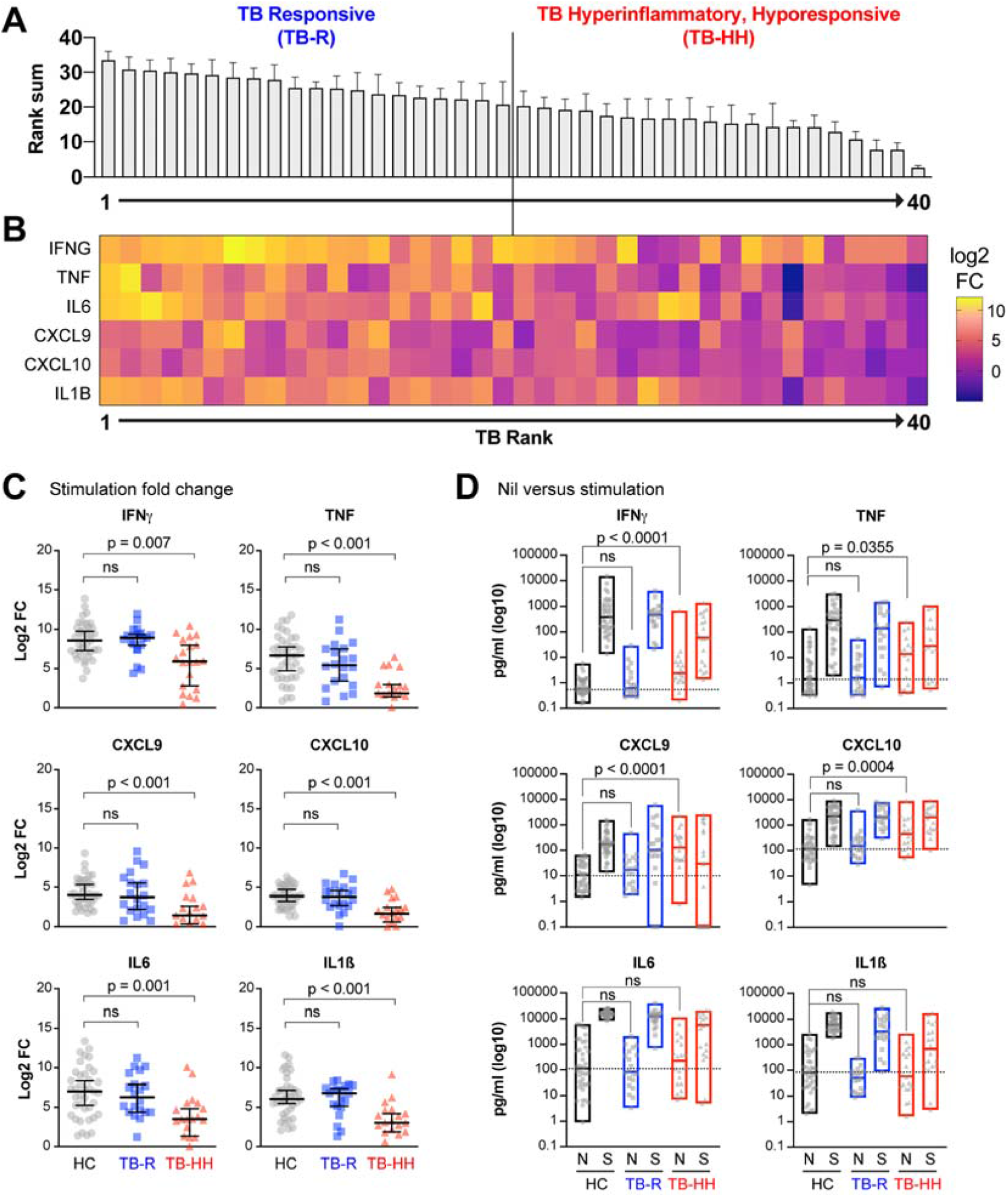
Identification of hyper-inflammatory, hyporesponsive cytokine production in TB patient endotypes. Whole blood from TB patients (n = 40) and healthy controls (n = 39) was stimulated overnight with or without mitogen (PHA), followed by measurement of cytokines and chemokines. (A) Samples were ranked for up-regulation of six cytokines to determine an overall rank sum (1 is lowest, with 40 highest). Using the rank sum value, TB patients were then split in half into “hypo-responsive” (red) and “responsive” (blue) groups. (B) Heatmap of cytokine expression as log2 fold change relative to controls. (C) Cytokine protein expression (log2 fold change) is graphed for each sub-group. Significance determined by Mann-Whitney non-parametric test. (D) Absolute protein expression with nil (N) or mitogen stimulation (S) expressed in pg/mL.

### Comparison of chemical compound signatures to TB endotype signatures

The Library of Integrated Network-based Cellular Signatures (LINCS) enables the identification and prioritization of putative drugs to treat a pathologic condition by countering its transcriptome signature. Comparing gene signatures for endotypes A and B to healthy controls, we obtained ranked lists of over 5,000 chemical compounds for each endotype and performed a comparative analysis. Previously identified candidates for host-directed therapy (HDT), including Vitamin D, glucocorticoids, non-steroidal anti-inflammatory drugs (NSAIDS), and retinoids, demonstrated connectivity scores that suggest a putative benefit for one endotype and either an inconsequential or contradictory response for the other endotype (Supplemental Figure 5). For example, HDAC inhibitors, such as vorinostat and phenylbutyrate, demonstrated transcriptomic signatures similar to endotype A, but dissimilar to endotype B.

## DISCUSSION

In the pre-antibiotic era, a fifth of humans with active tuberculosis survived more than 10-years[23], but present knowledge is inadequate to describe the underlying mechanisms of a sufficient immune response to overcome TB and contain *Mtb* infection. Further, to date, a single adjuvant host-directed therapy has not been identified, probably because the immune response to TB is protean and polymorphic. In this study, we identified clinically relevant TB endotypes by using unbiased clustering of unstimulated blood transcriptomes. Compared to controls, both endotypes displayed elevated gene expression related to pathways for inflammation and immunity, with higher levels among endotype A. In contrast, endotype B enriched for oxidative phosphorylation, the TCA cycle and pathways related to cellular proliferation, while endotype A demonstrated decreased pathways related to proliferation. Heme metabolism was upregulated in endotype A and downregulated in endotype B compared to controls, as described previously[6]. We derived a concise random forest classifier for TB endotypes, then used it to predicted endotypes in a validation cohort with richly annotated clinical outcomes; endotype A demonstrated slower times to bacterial clearance, and reduced incidence of disease cure.

Patients with TB are currently treated based on studies examining large heterogeneous groups. However, it is reasonable to hypothesize that subgroups exist within these large populations and that stratified and precision medicine strategies may improve outcomes[7]. These data provide support for individually stratified treatment approaches. Considering the animal model, *in vitro*, and human evidence, additional subtypes and endotypes will likely be identifiable when more robust epidemiology, strain characterizations, and functional immune analyses are integrated with transcriptomic results. It is notable that both endotypes exhibit elevated unstimulated gene expression levels of IFN-γ and TNF-□; however, in functional studies the TB patients with elevated basal IFN-γ and TNF-□ were less likely to upregulate IFN-γ and TNF-□ upon stimulation. Provided that future pair-wise transcriptomic and immune function studies confirm that endotype A displays characteristics of immune exhaustion (hyperinflammatory but hyporesponsive), this classification should guide future endotype-specific HDTs.

The trajectory analysis suggest that pathways related to immunity and inflammation monotonically progress from healthy controls to first endotype B and subsequently to endotype A. In contrast, cellular proliferation and oxidative phosphorylation and the TCA cycle increase in endotype B, but decrease in endotype A. Similarly, pathways related to proliferation decrease from controls to endotype A. This is a pattern similar to murine models of TB and other chronic infections[19, 24, 25] and therefore suggests that a stage-specific intervention can prevent the progression to terminal immune exhaustion in tuberculosis.

This study is limited in its capacity to determine appropriateness of the host immune response due to limited metadata and sub-optimal means to quantify bacillary burden. Immunity to *Mtb* is tightly regulated to avoid pathologic inflammation[2, 22, 26, 27]. Animal models have demonstrated that both IFN-γ and TNF□ require delicate homeostatic regulation with deficient responses allowing disease progression and exuberant responses resulting in immune-mediated pathology [2, 3, 22, 26, 28]. The included validation cohort used the best available measurement of bacillary burden (liquid culture time to positivity) and suggests that endotype A has a hyperinflammatory response with delayed culture conversion. Prospective studies need to combine gene expression analysis with functional immunology and quantitative measures of bacillary burden to clarify the appropriateness of host immunity. We anticipate that once endotypes are analyzed using robust multi-omic platforms, effective and pragmatic classifiers could use a minimal complement of informative features. Capitalizing on this minimized complement, cost-effective diagnostics could be developed and deployed at point-of-care in TB high burden settings.

The link between metabolism, particularly glycolysis, and immune function has been appreciated for over 90 years [20-22]. Metabolism is a mediator of the epigenetic mechanisms driving immune function[24, 25, 29-31]. Therefore, it is interesting that endotypes displayed incongruent regulation of genes and pathways related to metabolism, proliferation, and immune response. Many HDT candidates target these pathways. For example, metformin mediates the AKT-mTOR pathway, blunting cellular glycolysis and the TCA cycle leading to inhibition of chromatin conformational changes that drive antigen-induced immune function[29, 32]. Another inhibitor of mTOR, everolimus decreased TB-induced lung damage[27]. *In silico* evidence indicates the most pronounced benefit of everolimus for endotype A.

Previously identified candidates for HDTs include IFN-γ, GM-CSF, TNF□, TNF□ inhibitors, NSAIDS, Vitamin D, glucocorticoids, HDAC inhibitors, mTOR modulators, retinoids, and statins[27, 33]. The *in silico* analysis demonstrated that previously identified HDTs would perform better if applied in endotype-specific manners. If functional studies validate one endotype to have decreased immune responsiveness, then vitamin D or exogenous recombinant IFN-γ may be an appropriate HDT option. In contrast, if future validation studies demonstrate one endotype to have pathologic, exuberant immunity, then NSAID, TNF□ inhibitor, or glucocorticoid treatment would be appropriate. Animal and *in vitro* models that recapitulate the clinically relevant endotypes are also needed to better evaluate endotype-specific HDTs.

All included studies evaluated unstimulated host gene expression. TB is a chronic infection resulting in immune suppression. While many genes downstream of IFN-γ are elevated in TB patients at baseline[13, 15, 34, 35], they have decreased antigen-induced immune upregulation[36-39]. The multiplex ELISA data highlight the limitations of inferring immune function based on non-stimulated gene expression measurements; in fact, the group with elevated baseline cytokines (hyperinflammatory) was hyporesponsive upon stimulation. Additional gene expression subclusters were visible at higher resolution; however, biological relevance was not readily obvious in these sub-clusters. We speculate that the integration of transcriptomics with functional immune analysis, more robust epidemiology, and strain characterization will identify more than two endotypes.

Progression to TB is related to interactions among host, pathogen, and environmental factors. Progression to a specific endotype of TB is likely similarly related to as-yet unappreciated interactions. Unlike The Cancer Genome Atlas (TCGA), very limited epidemiology is available in existing public data repositories. Epidemiologic predispositions likely drive the divergent endotypes, including malnutrition, HIV, helminth, tobacco use, and/or indoor biomass fuel exposure. For example, despite successful deworming, previous schistosomiasis infection ablates mycobacterial immunity, leaving long-lasting immune suppression. We speculate that individuals with pre-existing immune suppression progress rapidly to endotype A, in contrast to previously healthy individuals.

In conclusion, this unbiased clustering provides additional evidence that there are multiple molecular host pathways modulated during TB [2-7]. This analysis of transcriptome and protein data from TB patients provides additional evidence for biologically distinctive TB endotypes that differentially affect clinical outcomes. Specifically, host gene expression in TB patient clusters into at least two endotypes with differential immune and metabolic transcriptomic signatures. These observations suggest that different endotypes display responses that are likely to have clinical and pathologic relevance and provides the basis for studies to evaluate endotype-specific host-directed therapies.

## Data Availability

All data are publicly available as per the manuscript.

---

**Supplemental Figure 1:**
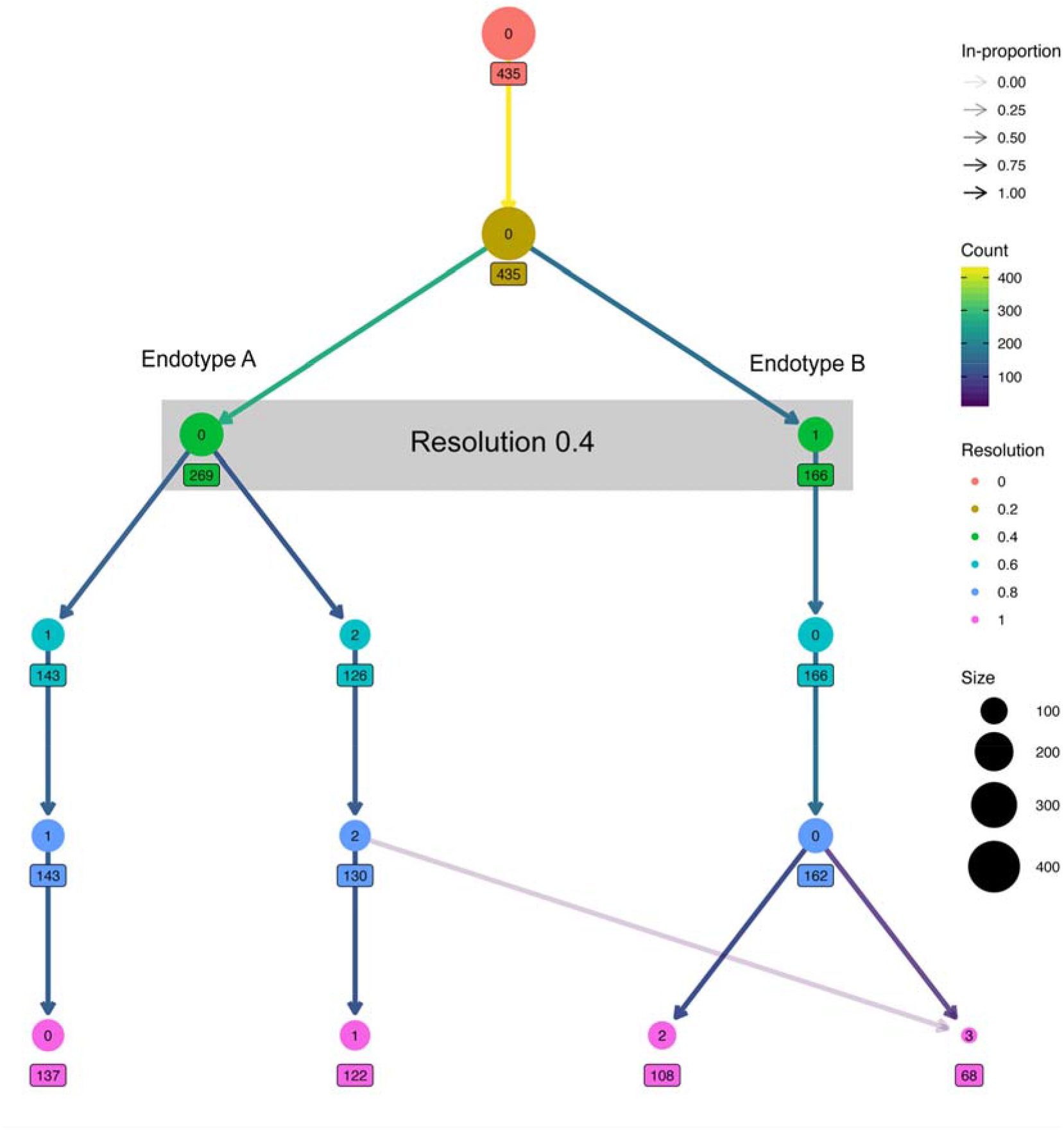
Hierarchical cluster tree identifying different clusters based on resolution.

**Supplemental Figure 2:**
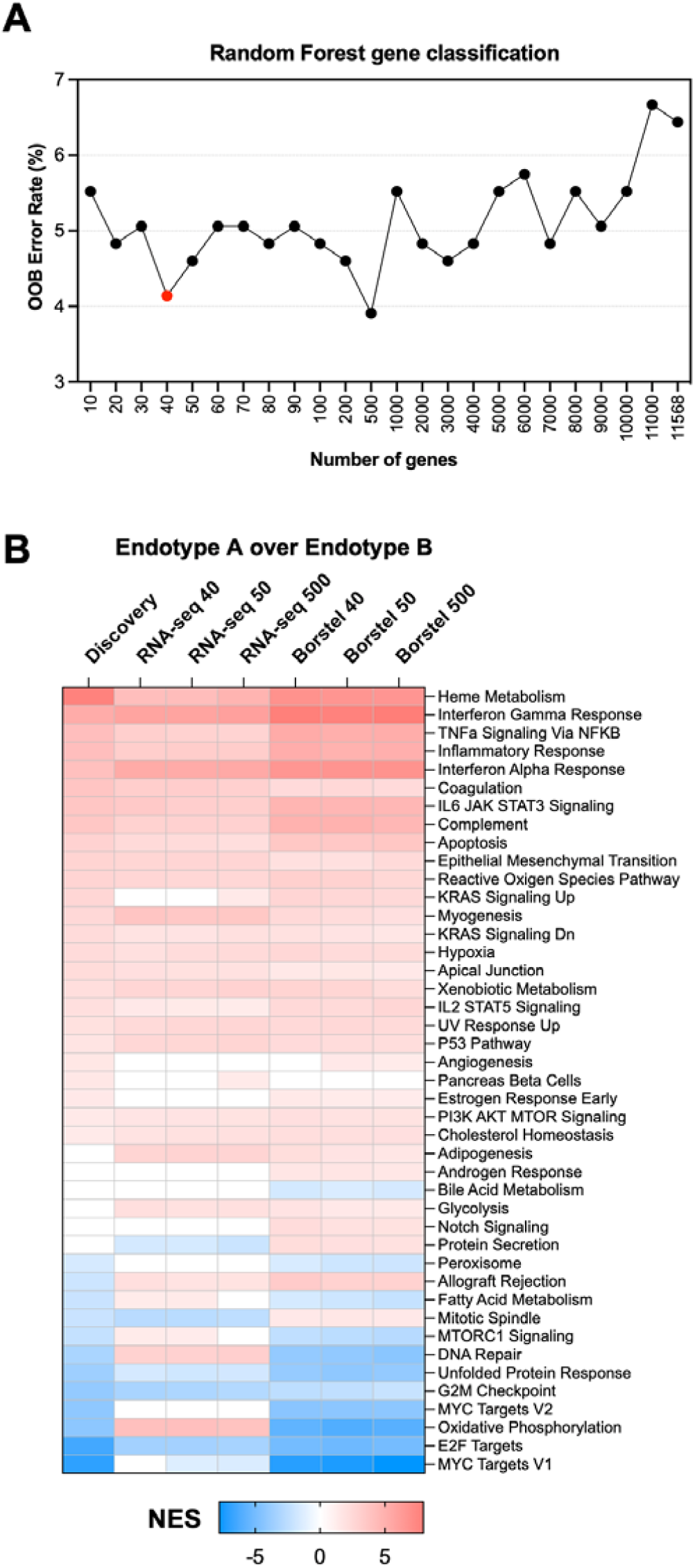
A) Out of bag error rate across a spectrum of genes for applying the Random Forest classifier. B) Heatmap of GSEA Normalized Enrichment Scores (NES) for Hallmark pathways using the 40, 50 and 500 gene TB endotype classifiers over discovery and validation cohorts.

**Supplemental Figure 3:**
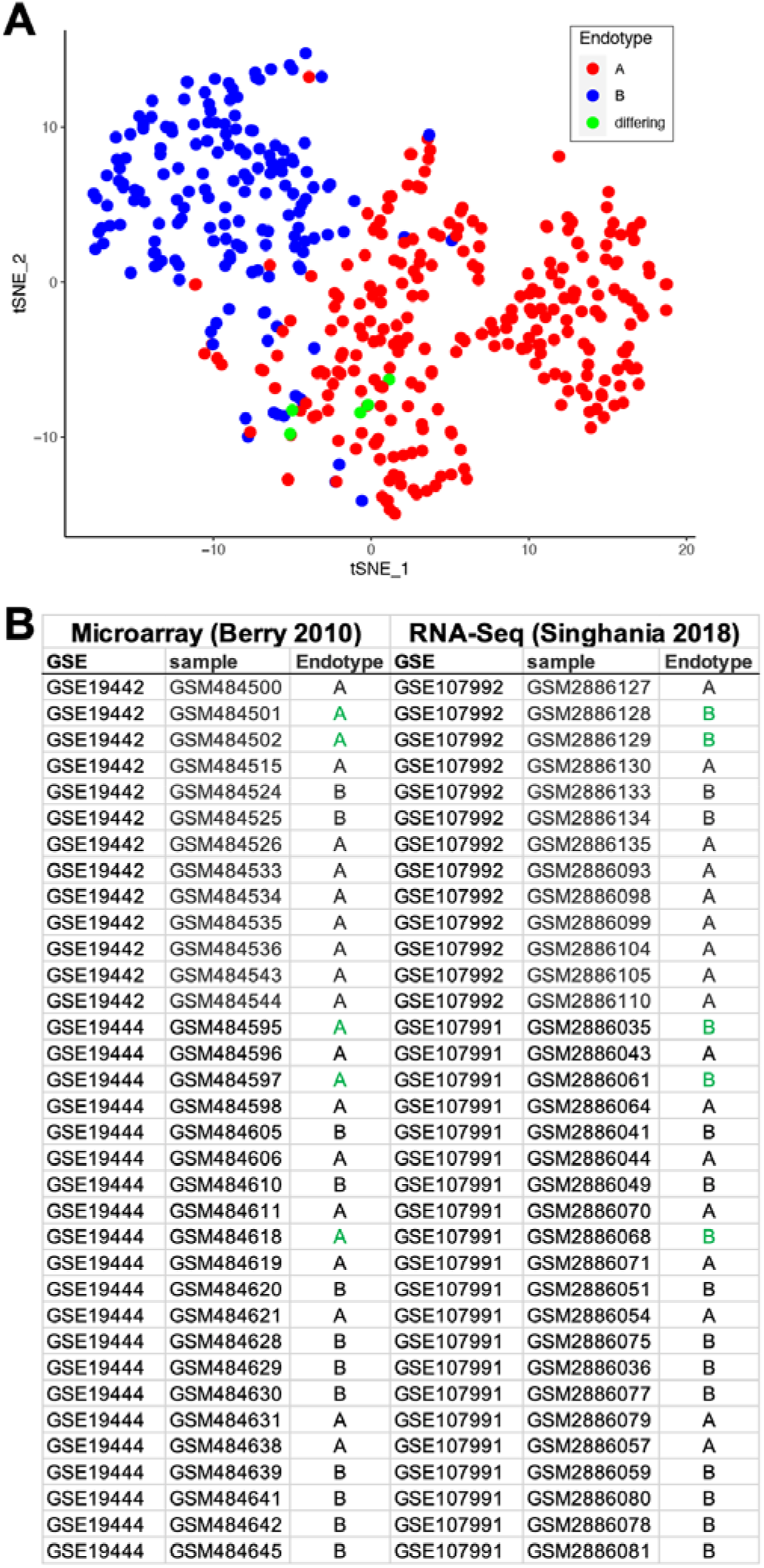
Concordance of sample classification for the same patient samples using microarray data in Berry 2010, compared to RNA-seq in Singhania 2018.

**Supplemental Figure 4.**
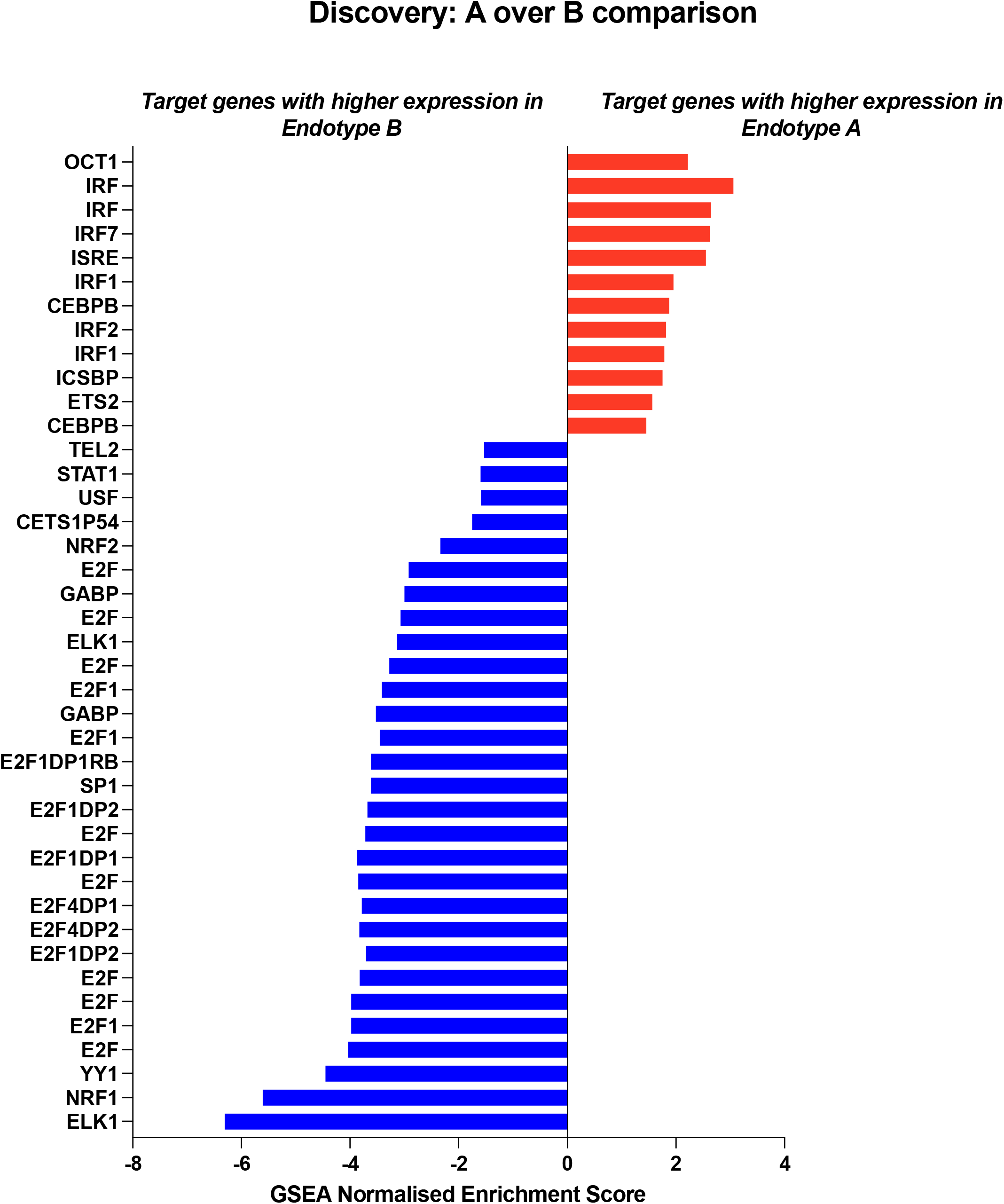
Differentially enriched Transcription Factor Targets pathways. Genes targeted by different transcription factors are divergent between endotype A and endotype B.

**Supplemental Figure 5.**
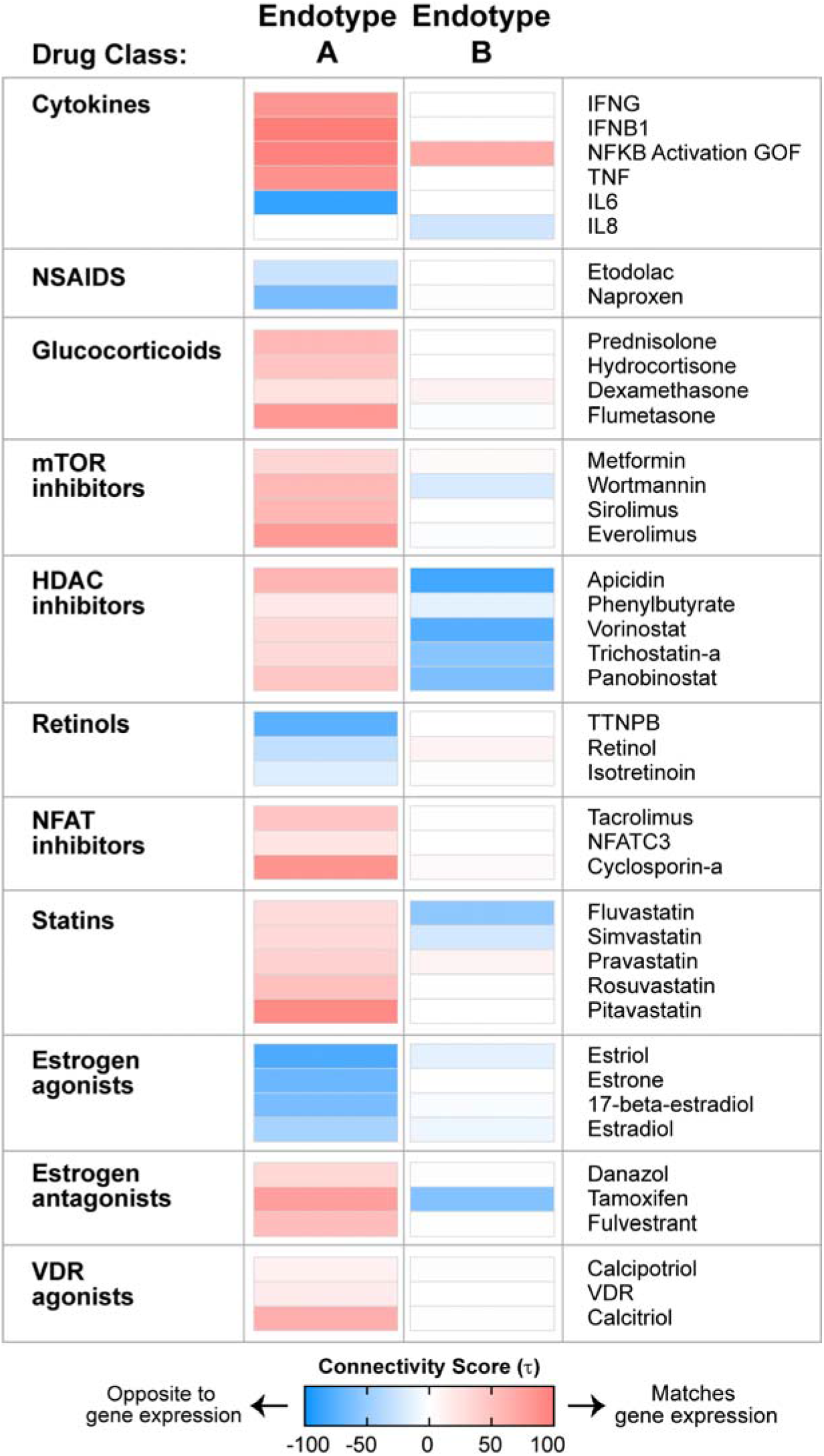
Heatmap of connectivity scores for select chemical compounds within the TB endotypes A and B based on the Library of Integrated Network-based Cellular Signatures (LINCS). Positive connectivity scores represent compounds inducing gene expression profiles similar to the endotype, while negative connectivity scores represent compounds inducing gene expression profiles antithetical to the endotype.

